# Increased Severity of New-onset Type 1 Diabetes in Children and Adolescents during the COVID-19 Pandemic: Experience from a Tertiary Care Center in Serbia

**DOI:** 10.1101/2023.11.20.23298741

**Authors:** Damjan Jovanovic, Jelena Blagojevic, Dejana Stanisavljevic, Maja Jesic

## Abstract

**Background and objectives:** Public health measures, parental fear of infection, and redeployment of medical resources in response to the COVID-19 pandemic might have led to a decrease in pediatric healthcare access. As a result, a delay in type 1 diabetes diagnosis might have occurred, leading to the worsening of its clinical presentation in the pediatric population. This study aimed to examine the clinical and biochemical features of new-onset DM1 in children and adolescents during the COVID-19 pandemic, comparing it to the pre-pandemic period.

**Materials and methods:** The clinical and biochemical features of diabetes observed during the COVID-19 period from April 1, 2020, until December 31, 2022, were compared with the period from April 1, 2017, until December 31, 2019. In the COVID-19 pandemic group, the clinical and biochemical features were compared between children with and without SARS-CoV-2 infection at diagnosis or before the diagnosis of DM1.

**Results:** During the COVID-19 pandemic, observed frequencies of DKA and severe DKA at diagnosis were 47.6% and 20.7%, both significantly higher than during the pre-pandemic period (an absolute increase of 15% and 11.3%, respectively). In the COVID-19 group, blood pH levels were significantly lower than in the pre-pandemic group, while HbA1c levels were higher. Clinical and biochemical features of diabetes in children with SARS-CoV-2 infection at or before the diagnosis were not significantly different compared to children without an infection.

**Conclusion:** We report a significant worsening of the clinical presentation of new-onset type 1 diabetes and an increase in the frequency of DKA and severe DKA at diagnosis during the COVID-19 pandemic. Further studies are necessary to gain quantitative insight into pediatric healthcare availability in Serbia.

## 1. Introduction

COVID-19 is an infectious disease caused by Severe Acute Respiratory Syndrome Coronavirus 2 (SARS-CoV-2). The first cases of SARS-CoV-2 infection were reported in December 2019 in Wuhan, China. [1] The first laboratory-confirmed case of SARS-CoV-2 infection in Serbia was reported on March 6, 2020 [2], and the World Health Organization (WHO) declared COVID-19 a pandemic on March 11 of the same year. [1] The COVID-19 pandemic has proven itself to be a great challenge for healthcare systems and societies as a whole throughout the world. In the Republic of Serbia, as in many other countries, a series of public health measures were introduced to curb the viral spread throughout the population. In response to a rising number of patients with a moderate to severe clinical presentation of the SARS-CoV-2 infection, a substantial number of hospitals were adapted for the treatment and care of COVID-19. Additionally, numerous healthcare professionals were redeployed from their regular units to specialized COVID-19 centers. While severe forms of the disease in children are uncommon, it’s essential to recognize that poor outcomes are still plausible within the pediatric population. [3,4] Furthermore, children often act as asymptomatic carriers of the virus and may serve as a significant link in the transmission chain of the disease. [5,6] For those reasons, children were also subjected to public health measures.

Public health measures, parental fear of infection, and redeployment of healthcare system resources may have led to a decrease in pediatric healthcare availability and resulted in a delay in the diagnosis and treatment of other diseases [7–9], such as type 1 diabetes. A survey of the perception and experience of healthcare professionals from pediatric centers carried out during the first year of the pandemic showed that a significant number of respondents reported a delay in the diagnosis of diabetes and an increase in DKA frequency. [10] Indeed, a number of groups reported an increase in DKA and severe DKA prevalence on the initial presentation of newly diagnosed type 1 diabetes in children and adolescents during the pandemic. [11–15] Healthcare system overload and a decrease in pediatric healthcare access and utilization may have led to a delay in DM 1 diagnosis and a worsening of the clinical picture and biochemical parameters of the disease in the Republic of Serbia as well. To evaluate this hypothesis, we collected the data on disease characteristics (blood glucose levels, HbA1c, pH, DKA frequency and severity) of newly diagnosed DM 1 cases presenting to the University Children’s Hospital, Belgrade, Serbia, over the 33 months of the COVID-19 pandemic and compared them with the data for the 33-month period that immediately preceded the pandemic. Besides the indirect effects of the pandemic on the clinical picture of DM 1, there is a possibility of a direct effect of the SARS-CoV-2 infection on the biology of the disease. To evaluate the mechanistic effect of infection on the severity of metabolic disorder in DM 1, we compared clinical and biochemical features of the disease between children who had the SARS-CoV-2 infection at or before the diagnosis of DM 1 and children who didn’t.

## 2. Materials and Methods

The study was designed as a single-center, retrospective observational study. The study population included children and adolescents less than 18 years old presenting to the Department of Pediatric Endocrinology at the University Children’s Hospital (Belgrade, Serbia) with new-onset type 1 diabetes. The COVID-19 pandemic group included children and adolescents admitted to our department with a new diagnosis of DM 1 from April 1, 2020, to December 31, 2022, while the control group was comprised of patients newly diagnosed with type 1 diabetes in the period from April 1, 2017, to December 31, 2019. During the COVID-19 pandemic, every child was tested for SARS-COV-2 upon admission to the hospital using a rapid antigen test (RAT), and positive results were verified using polymerase chain reaction (PCR).

DM1 was defined according to the criteria of the International Society for Pediatric and Adolescent Diabetes (ISPAD). [16] Patients admitted with previous type 1 diabetes, non-DM1, or unclear types of diabetes were excluded from the study. Data collected from paper and electronic medical charts included sex, age, residence, date of DM 1 diagnosis, as well as the clinical features of the disease (symptom duration, blood pH, blood glucose levels, and DKA/severe DKA on presentation). Diabetic ketoacidosis (DKA) was defined as the presence of: (1) blood glucose > 11 mmol/l, (2) blood pH < 7.3 or bicarbonate < 15 mmol/l, and (3) ketonemia or ketonuria. Severe DKA was defined as pH < 7.1. [17] In the pandemic group, data were collected on the presence of SARS-CoV-2 infection before the DM1 diagnosis (history of a positive PCR, positive RAT in the presence of symptoms, or presence of SARS-CoV-2 IgM/IgG antibodies at any point before the diagnosis of type 1 diabetes) or at the initial presentation of DM1 (results of RAT upon admission to the hospital).

### 2.1. Statistical analysis

Data are presented as means ± standard deviation for continuous variables with a normal distribution or as numbers (percentages) for nominal variables. For comparisons between the pandemic group and pre-pandemic control group, as well as between the children with and without SARS-CoV-2 infection before or at the diagnosis of DM1 in the pandemic group, t-test and ANOVA were conducted for continuous variables, and the Chi-square test for nominal variables. A two-sided P < 0.05 was considered statistically significant. The data were analyzed using the Statistical Package for the Social Sciences (SPSS) Version 21 software.

## 3. Results

A total of 143 children and adolescents presented with a new-onset DM1 over the 33-month study period during the COVID-19 pandemic, in comparison to 135 patients presenting during the equally lasting control period preceding the pandemic. The demographic characteristics of the pandemic and control groups did not differ significantly (Table 1). The average number of new DM1 cases per month during the pandemic and control period was 4.33 and 4.09, respectively. The highest average number of cases per month was recorded in 2021 (5.33), while the lowest was observed in 2018 (3.00) (Figure S1A).

**Table 1.**
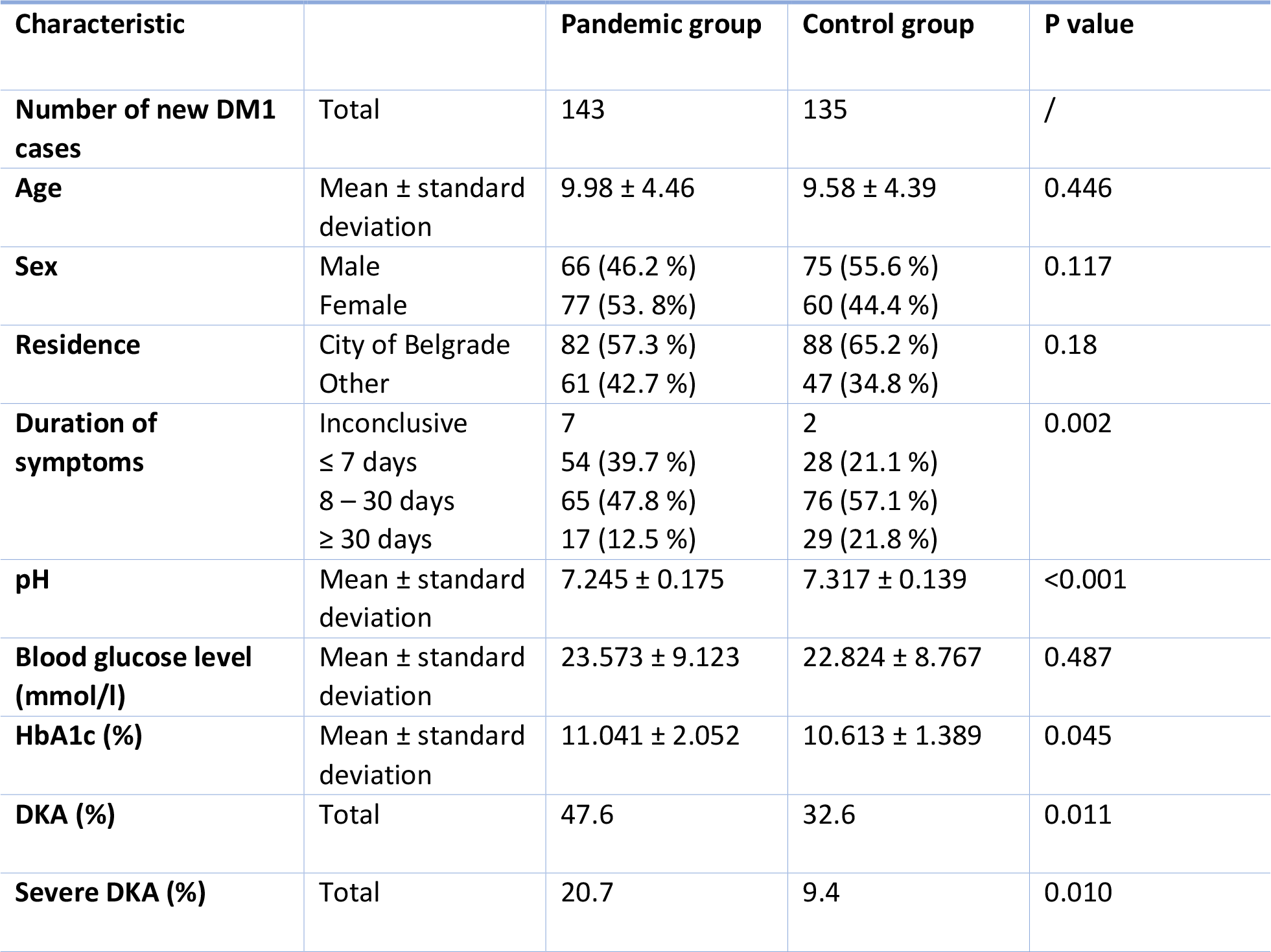
Demographic and clinical/biochemical characteristics of new onset DM1 during the COVID-19 pandemic period (April 1, 2020 to December 31, 2022) and the pre-pandemic control group (April 1, 2017 to December 31, 2019) and the pandemic group.

Mean blood pH values were significantly lower in the pandemic group compared to the control group (7.245 ± 0.175 during the pandemic vs. 7.317 ± 0.139 during the control period, P < 0.001) (Figure 1B), while the HbA1c levels were significantly higher (11.041 ± 2.052 vs. 10.613 ± 1.389, P = 0.045) (Figure 1C). Mean blood glucose values were higher in the pandemic group, albeit non-significantly. The percentage of children presenting within 7 days of the onset of symptoms was higher during the pandemic (39.7% vs. 21.1%, P = 0.002), while the percentage of children with the duration of symptoms > 30 days was lower (12.5% vs. 21.8%) (Table 1). During the COVID-19 pandemic, the frequency of DKA at the initial presentation of type 1 diabetes was strikingly increased as compared to the control period (47.6% during the pandemic vs. 32.6% during the control period (P = 0.011), an absolute increase of 15%). The presence of severe DKA at the onset of DM1 was also significantly more common in the pandemic group (20.7% vs. 9.4%, P = 0.01), with an absolute increase of 11.3%. (Table 1 and Figure 1A) pandemic group, higher blood glucose levels were noticed in younger patients (27.096 ± 7.322 in children aged < 5 years, 23.832 ± 9.402 in children aged 5–11 years, and 21.928 ± 9.243 in children aged ≥ 11 years, P = 0.049). Such a trend of age-related decrease in mean blood glucose level wasn’t seen in the control group (Table 2 and Figure S2). The duration of symptoms in the pre-pandemic period was significantly shorter among children aged < 5 years in comparison to other age categories (Table 2). There were no significant differences in the duration of symptoms between the age groups during the COVID-19 pandemic. In the control group, the frequency of DKA was the highest in children aged < 5 years. While the observed difference did not reach statistical significance, it’s worth noting that the p-value (0.056) was borderline, and its significance would have likely been revealed in a larger sample. (Table 2)

**Table 2.**
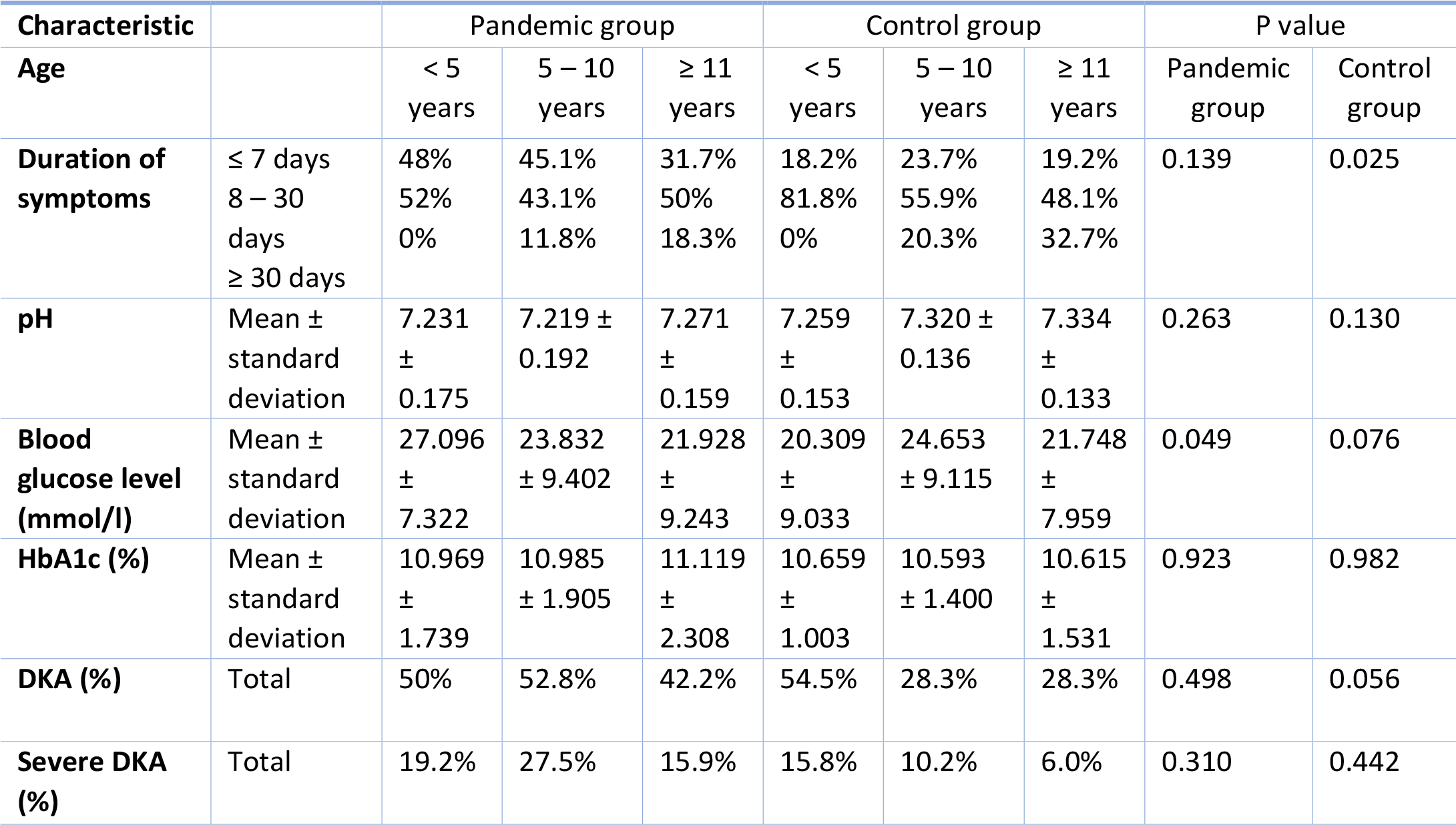
Age-specific distribution of clinical and biochemical characteristics in individuals newly diagnosed with DM1 during the COVID-19 pandemic and the control period.

**Figure 2.**
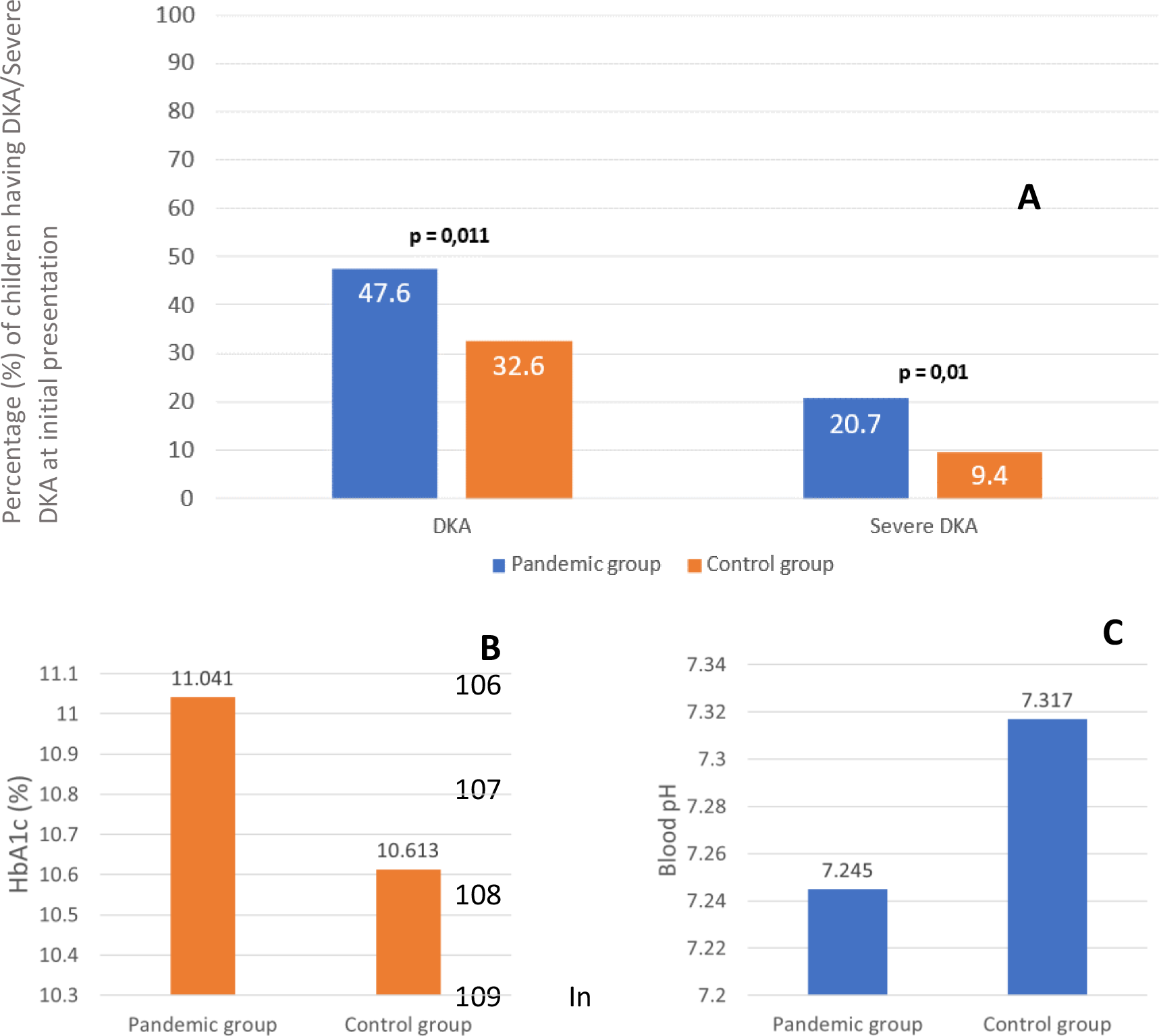
Clinical and biochemical characteristics of new onset DM1 during the COVID-19 pandemic period (April 1, 2020 to December 31, 2022) and the pre-pandemic control group (April 1, 2017 to December 31, 2019) and the pandemic group. **A**. During the COVID-19 pandemic frequencies of DKA and severe DKA at diagnosis were 47.6% and 20.7%, both significantly higher than during the pre-pandemic period (an absolute increase of 15% and 11.3%, respectively). **B, C**. HbA1c levels were significantly higher in the pandemic group, while blood pH levels were higher.

A total of 14 children (9.8%) from the pandemic cohort tested positive for SAR-CoV-2 upon admission (N = 6, 36.2%) or had a positive history of a laboratory-confirmed SARS-CoV-2 infection before the type 1 diabetes diagnosis (N = 8, 62.8%). All children were either asymptomatic or had mild symptoms of COVID-19. An additional 16 patients (11.18%) had a history of COVID-19 symptoms or a close contact (household member with a confirmed SARS-CoV-2 infection) during the pandemic. However, no definitive confirmation of the infection was established (positive PCR, positive RAT with symptoms, or the presence of SARS-CoV-2 IgM/IgG antibodies) at any point before the diagnosis of DM1. Frequencies of DKA and severe DKA at the onset of DM1 were similar between children who didn’t have a SARS-CoV-2 infection before the diagnosis of DM1 and children who tested positive for SARS-CoV-2 upon admission or had a positive history of a laboratory-confirmed SARS-CoV-2 infection before the type 1 diabetes diagnosis (Table 3).

**Table 3.** Characteristics of new onset DM1 in children with and without SARS-CoV-2 infection before or at diagnosis of diabetes.

| Characteristic |  | No SARS-CoV-2 infection | SARS-CoV-2 infection | P value |
| --- | --- | --- | --- | --- |
| **Number of new DM1 cases** | Total | 129 (91.2%) | 14 (9.8%) |  |
| **Age** | Mean ± standard deviation | 9.713 ± 4.388 | 12.450 ± 4.514 | 0.396 |
| **Sex** | Male<br>Female | 63 (48.8%)<br>66 (51.2%) | 3 (21.4%)<br>11 (78.6%) | 0.051 |
| **Residence** | City of Belgrade<br>Other | 75 (58.1%)<br>54 (41.9 %) | 7 (50%)<br>7 (50%) | 0.559 |
| **Duration of symptoms** | ≤ 7 days<br>8 – 30 days<br>≥ 30 days | 39.7%<br>47.8%<br>12.5% | 57.1%<br>28.6%<br>14.3% | 0.295 |
| **pH** | Mean ± standard deviation | 7.239 ± 0.179 | 7.296 ± 0.130 | 0.254 |
| **Blood glucose level (mmol/l)** | Mean ± standard deviation | 23.676 ± 8.814 | 22.629 ± 11.964 | 0.685 |
| **HbA1c (%)** | Mean ± standard deviation | 11.128 ± 1.992 | 10.250 ± 2.477 | 0.129 |
| **DKA (%)** | Total | 47.3 | 50.0 | 0.847 |
| **Severe DKA (%)** | Total | 22.2 | 7.1 | 0.187 |

## 4. Discussion

Lower mean blood pH, higher mean HbA1c levels, and an increased prevalence of DKA/severe DKA on the initial presentation of new-onset DM1 in the period from April 2020 to December 2022 in comparison to the control period point to the worsening of the clinical picture of newly diagnosed type 1 diabetes among children and adolescents during the pandemic in Serbia. Our results correspond to findings reported by a number of groups from other countries [11–14,18–20], as well as several meta-analyses. [21,22] Multiple factors have likely contributed to the substantial worsening of the clinical picture and biochemical parameters of new-onset type 1 diabetes. In Serbia, as in numerous other countries, a series of public health measures were introduced to suppress the transmission of SARS-CoV-2 in the population and flatten the epidemic curve. As children may serve as asymptomatic carriers of the virus and less commonly develop severe disease, public health interventions were implemented for the pediatric population as well. Recommendations to decrease contact with other individuals were issued, the lockdown was imposed several times, many people had to work from home, and schools and most other public institutions were closed. Healthcare system overload led to changes in the organization of healthcare services. In response to a high influx of moderate and severe cases of SARS-CoV-2 infection that required hospitalization, a significant portion of healthcare professionals were redeployed from their regular units to centers adapted for the treatment and care of patients with COVID-19, and scheduling specialist appointments became increasingly difficult. Public health measures, parental fear of infection, healthcare system overload, and a decrease in pediatric healthcare availability and utilization may have led to a delay in the diagnosis and treatment of newly diagnosed DM1 in children and adolescents. Earlier investigations have demonstrated that DKA is a preventable complication and that its risk is increased by a delay in DM1 diagnosis. [23,24] Moreover, the findings of the increased prevalences of DKA and severe DKA at the onset of type 1 diabetes are particularly concerning, given that diabetic ketoacidosis is a life-threatening acute complication of diabetes and commonly requires admission to the PICU unit.

Recent findings from several studies give evidence of an increased incidence of pediatric DM1 during the COVID-19 pandemic [21,22,25,26]. Viral infections are well recognized as potential triggers for immune-mediated destruction of pancreatic β cells in type 1 diabetes, and previous epidemiological research has shown a rise in new cases of DM1 following viral epidemics [27]. SARS-CoV-2 was recently identified as a risk factor for the development of islet autoimmunity in young children with an increased genetic risk of DM1. [28] The number of newly diagnosed DM1 cases was similar between groups in this study. However, even though the University Children’s Hospital in Belgrade is a tertiary care center and admits the most newly diagnosed type 1 cases in Serbia, it is not the only center to provide care for pediatric diabetes in the country. As we don’t have insight into the complete number of new type 1 diabetes cases diagnosed in the territory of Serbia during the observation period, we can’t estimate DM1 incidence during the pandemic. Previously, an increase in the incidence of DM1 during the pandemic was reported in a multi-center study in Vojvodina, which is an autonomous province of the Republic of Serbia. [29] In that study, Vorgucin et al. reported that the incidence of DM1 in 2021 was the highest in a five-year period from 2017 to 2021. That corresponds with the finding from our study that the year 2021 had the highest average number of cases per month, with the average number of cases per month decreasing to the near pre-pandemic level in 2022.

Some authors point out the possibility of a direct relationship between SARS-CoV-2 infection and a more severe clinical picture and DKA at the initial presentation of DM1. (18) SARS-CoV-2 penetrates the cells after binding to the cell surface receptor angiotensin-converting enzyme 2 (ACE2) and proteolytic cleavage of the viral spike (S) protein by transmembrane serine protease 2 (TMPRSS2) [30]. ACE 2 and other viral entry factors (TMPRSS2, neuropilin 1, transferrin receptor, and furin) are expressed on the surface of multiple pancreatic cell types [31], and it was demonstrated that SARS-CoV-2 can infect and replicate in β-cells of Langerhans islands [32–35], leading to β-cell transdifferentiation [34], suppression of insulin secretion, and induction of apoptotic pathways [35]. However, the significance of the SARS-CoV-2 mechanistic contribution to pancreatic islet dysfunction and the severity of metabolic disturbance in DM1 is not yet clearly established *in vivo*. In this study, a significant difference in clinical and laboratory parameters of disease severity in DM1 between children with and without SARS-CoV-2 infection before or at DM1 onset was not seen. Therefore, we believe that the worsening of clinical and laboratory indicators of disease severity in children was primarily caused by the indirect effects of the pandemic on pediatric healthcare service organization and access. Nevertheless, the number of SARS-CoV-2-positive children in our study was small, consistent with the epidemiology of SARS-CoV-2 infection in the pediatric population. Additional studies are necessary to more precisely evaluate the direct relationship between viral infection and the severity of new type 1 diabetes at diagnosis.

This study has several important limitations. The study is single-center and conducted on a relatively small sample. A larger multi-center study would provide a better insight into the effects of the pandemic on pediatric DM 1 in the territory of Serbia. On the other hand, several strengths compensate for these limitations. The major strength of this study is its long observation period, which has covered almost the entire course of the pandemic in Serbia. To our knowledge, no similar study so far has had an observation period longer than 24 months. The study is also characterized by the systematic collection of data on the presence of SARS-CoV-2 infection and provides an opportunity to investigate the direct effects of the viral infection on the severity of DM1.

## 5. Conclusions

In conclusion, our research adds to the growing body of literature on the relationship between the severity of new-onset DM1 in children and the COVID-19 pandemic. An increase in DKA and severe DKA frequencies at the onset of type 1 diabetes, lower mean blood pH values, and higher mean HbA1c levels confirm that the COVID-19 pandemic was associated with an increase in clinical and laboratory parameters of severity of metabolic disorder in newly diagnosed cases of DM1 in the pediatric population in Serbia. We believe that the worsening of the clinical picture of type 1 diabetes and the increase in DKA/severe DKA at onset were caused by the interaction of multiple factors, among which healthcare system overload and a decrease in pediatric healthcare access and utilization stand out as the most important. Further studies are necessary to gain quantitative insight into pediatric healthcare availability and access in the Republic of Serbia.

## Author Contributions

Conceptualization, D.J. and M.J.; methodology, D.J., M.J., and D.S.; software, D.S.; validation, D.J., M.J., and D.S.; formal analysis, D.S. and D.J.; investigation, D.J. and J.B.; resources, M.J.; data curation, D.J., and J.B; writing—original draft preparation, D.J.; writing—review and editing, D.J., M.J., and D.S.; visualization, D.J. and D.S.; supervision, M.J.; project administration, M.J.; funding acquisition, M.J. All authors have read and agreed to the published version of the manuscript.

## Funding

This research received no external funding

## Institutional Review Board Statement

The study was conducted in accordance with the Declaration of Helsinki, and approved by the Ethics Committee of the University Children’s Hospital, Belgrade (protocol code 130-26/21, 2021).

## Informed Consent Statement

Informed consent was obtained from all subjects or their parents or guardians involved in the study for admission to hospitals and the procedures performed during hospitalization. A waiver was approved for individual written informed consent to publish this paper based on non-identifiable use of previously obtained retrospectively collected data.

## Data Availability Statement

Data is unavailable due to privacy and ethical restrictions

## Acknowledgments

The authors would like to acknowledge the children and families cared for by the University Children’s Hospital.

## Conflicts of Interest

The authors declare no conflict of interest.

## Supplementary Materials

**Figure S1.**
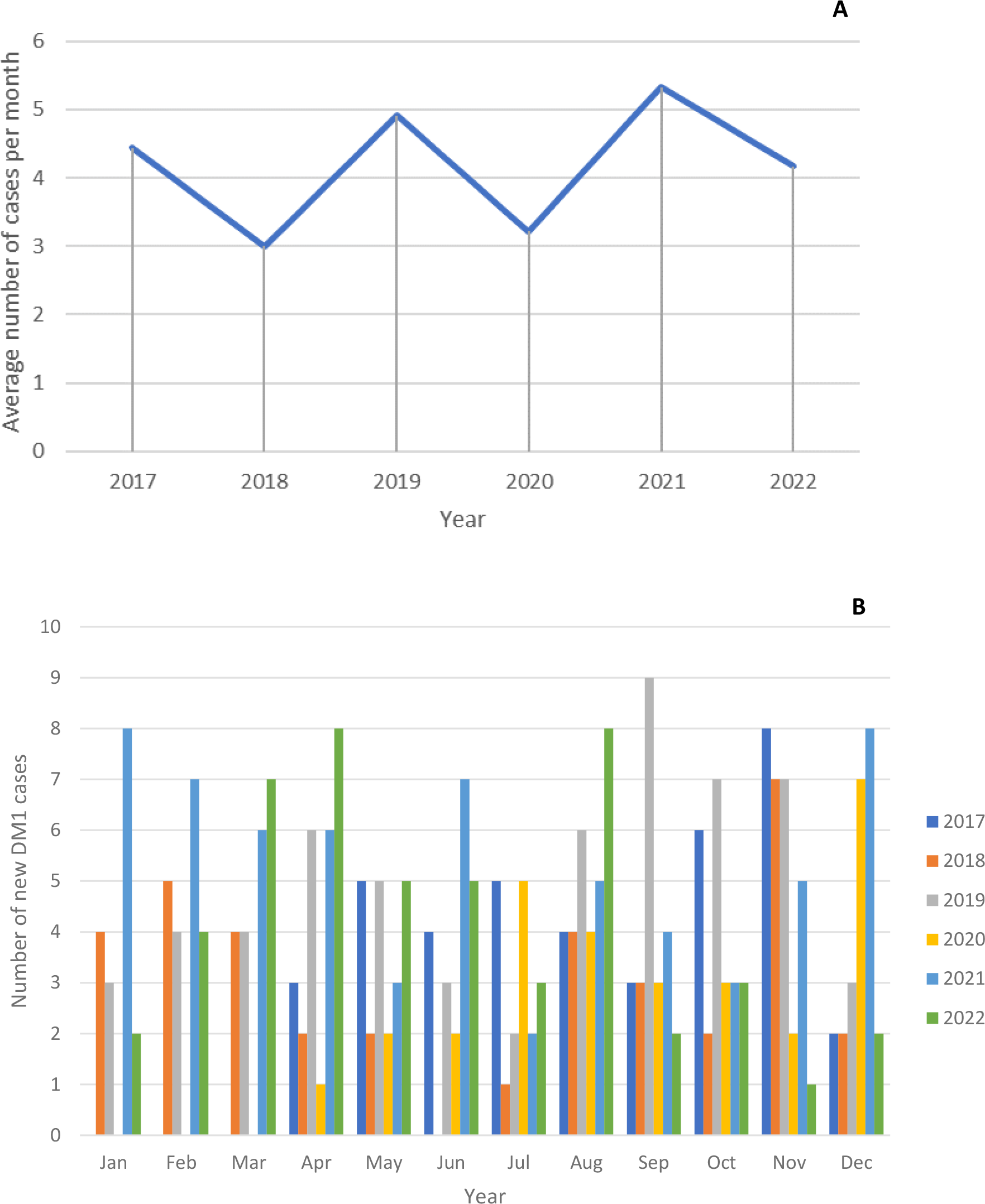
**A**. Average number of cases per month for every year. **B**. Total number of monthly cases for every year

**Figure S2.**
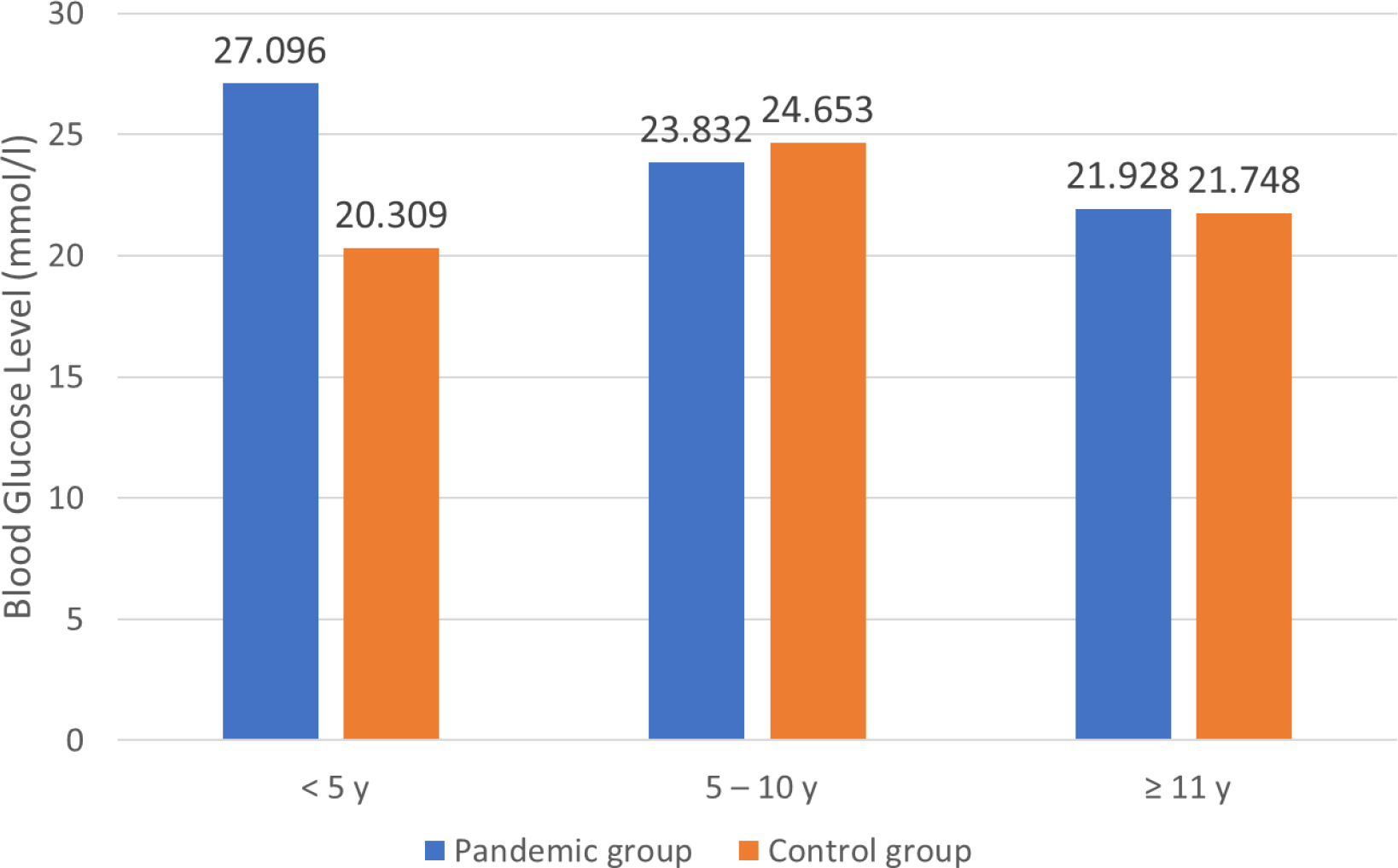
In the pandemic group, higher blood glucose levels were noticed in younger patients (27.096 ± 7.322 in children aged < 5 years, 23.832 ± 9.402 in children aged 5–11 years, and 21.928 ± 9.243 in children aged ≥ 11 years, P = 0.049). Blood glucose level differences among age categories weren’t significant in the control group (p = 0.076)

## Notes

### Competing Interest Statement

The authors have declared no competing interest.

### Funding Statement

This study did not receive any funding

### Author Declarations

Ethics Committee of the University Children's Hospital, Belgrade gave ethical approval for this work (protocol code 130-26/21)

